# Identification of Potential Blood-based Biomarkers for Multiple Sclerosis

**DOI:** 10.1101/2022.08.08.22278573

**Authors:** Adrija Kundu

## Abstract

Multiple Sclerosis (MS) is a disease affecting millions of people worldwide caused by an abnormal immune response that deteriorates the myelin sheath encapsulating neurons, drastically impairing neural communication. Currently, no tests can guarantee the diagnosis of MS until symptoms become severe. Early-stage diagnosis for MS is beneficial as it allows for an early start to treatment that slows its progression. This project aimed to analyze data from gene expression studies and identify commonly differentially expressed genes (DEGs) in the blood of MS patients. 183 overexpressed DEGs were identified and a diagram depicting protein interactions was generated with STRINGDB. Cytoscape was used to analyze interactions and recognize Hub genes. Further analysis on PANTHER showed that the biological processes of “tRNA aminoacylation for protein translation” and “mitochondrial translational elongation” were enriched in the upregulated DEGs. Furthermore, processes directly connected to the immune system such as the “Fc receptor signaling pathway” and “antigen processing and presentation of peptide antigen” were overrepresented. The identified Hub genes RPS27A, NSA2, RPS15A, and POLR2C are potential biomarkers for MS and warrant further study. Overall, this study could provide greater insight into the molecular pathways of MS and evidence to support which genes are overexpressed in MS.

## Introduction

Currently, an estimated 2.8 million people worldwide suffer from MS and its prevalence has increased in every world region since 2013. [1]. The disease is mediated by autoreactive lymphocytes that enter the Central Nervous System upon crossing the blood-brain barrier and cause demyelination, gliotic scarring, and axonal loss, ultimately creating lesions in the brain[2]. The symptoms vary based on where the lesions occur. For example, cerebellum lesions can cause vertigo whereas lesions in the brainstem can cause diplopia, commonly called double vision[3]. By the time patients develop obvious symptoms of MS, the disease has progressed to its more severe stage. While there is no cure for MS, there are treatment procedures that lessen the severity of symptoms and make it easier to recover from attacks. However, these treatment procedures are most effective when MS is detected at its early stages. However, this is a challenge because confirmatory symptoms don’t show up until the late stages.

Therefore, identifying biomarkers could prove very beneficial as they’d allow for early-stage detection of MS. Currently, there are numerous potential mediums for biomarker discovery such as blood and saliva[4]. However, as no definitive biomarkers have been identified, MRI scans and Cerebrospinal Fluid Taps are used by doctors to confirm MS. These procedures are both more dangerous and costly in comparison to blood tests, for example.

Identification of blood-based biomarkers would not only allow for the noninvasive detection of MS at early stages but could also be applied to develop therapeutics that could significantly slow down the progression of MS.

RNA-seq and microarrays can be used to profile blood-based biomarkers [5]. These methods are used in gene expression studies. Differential gene expression between MS patients and healthy controls could provide greater insight into the molecular pathways of MS and also aid in identifying biomarkers for diagnosis.

In this study, microarray data from three separate studies by different authors were analyzed to identify potential biomarkers for early-stage MS. Each of the studies provided gene expression data corresponding to the blood of MS patients and healthy controls. Three separate studies were obtained because sometimes, there may be discrepancies between studies due to varied methodologies, sample characteristics, storage methods, etc. [6-7] Therefore, identifying biomarkers common to multiple studies allows for greater confidence. In this present study, the genes differentially expressed across the three datasets were identified and further analyzed.

## Methods

### Obtaining Datasets for Analysis

The datasets analyzed in this project were obtained from the Gene Expression Omnibus (GEO), an online repository managed by NCBI that provides public expression profiling and RNA methylation profiling data [8-9]. The following keyphrases were used to search for the data sets on GEO: “multiple sclerosis”, “blood”, “homo sapiens”, and “expression profiling by array”. The datasets chosen for this study were GSE17048, GSE21942, and GSE23832[10-12]. Across the three datasets, there were a total of 122 samples with 58 MS Samples and 64 control samples. The MS samples across the three datasets were of patients with clinically definite MS, meaning that they had occurrences of remissions and at least 2 relapses with each lasting more than 24 hours and occurring more than one month apart. The expression profiling for each of the datasets was done by obtaining RNA from blood. mRNA was then isolated from the other types of RNA and then complementary DNA (cDNA) was generated and amplified through Reverse Transcriptase Polymerase Chain Reaction (RTPCR). Then, expression data was collected on microarrays based on the hybridization of the ssDNA probes on the microarrays by the cDNA[13].

**Table 1:**
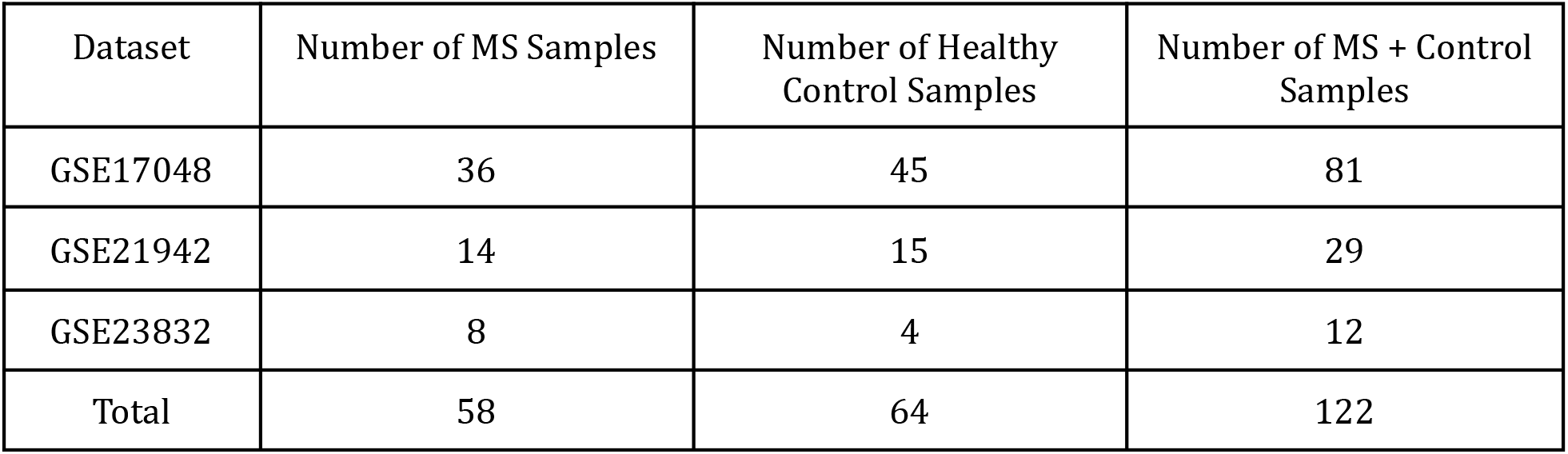
While the entirety of GSE21942 and GSE23832 reflected the clinically definite MS conditions, only 36/99 MS Patient samples in GSE17048 reflected these conditions and thus the remaining 63 were excluded from the study.

| Dataset | Number of MS Samples | Number of Healthy Control Samples | Number of MS + Control Samples |
| --- | --- | --- | --- |
| GSE17048 | 36 | 45 | 81 |
| GSE21942 | 14 | 15 | 29 |
| GSE23832 | 8 | 4 | 12 |
| Total | 58 | 64 | 122 |

### Differential Expression Analysis

GEO2R, provided on the GEO Databus, was used to obtain the distribution of gene expression values[8]. The tool is a browser-based software that provides a list of statistically significant DEGs between two groups (In this scenario, MS Patients & Healthy Controls). Each dataset had thousands of statistically significant DEGs. GEO2R also calculated the Fold Change Values (Ratio of average expression in experimental group to average expression in control group) and p values (obtained from T-Tests). No outliers were identified in any of the three data sets. This data was downloaded and imported to Google Sheets where the DEGs with a p-value greater than 0.05 were removed from the list. Following this, the DEGs were sorted from highest Fold Change (Most overexpressed in MS sample) to lowest Fold Change (Most underexpressed in MS sample). Only the genes with a fold change value greater than 0 were kept in the list.

**Figure 1:**
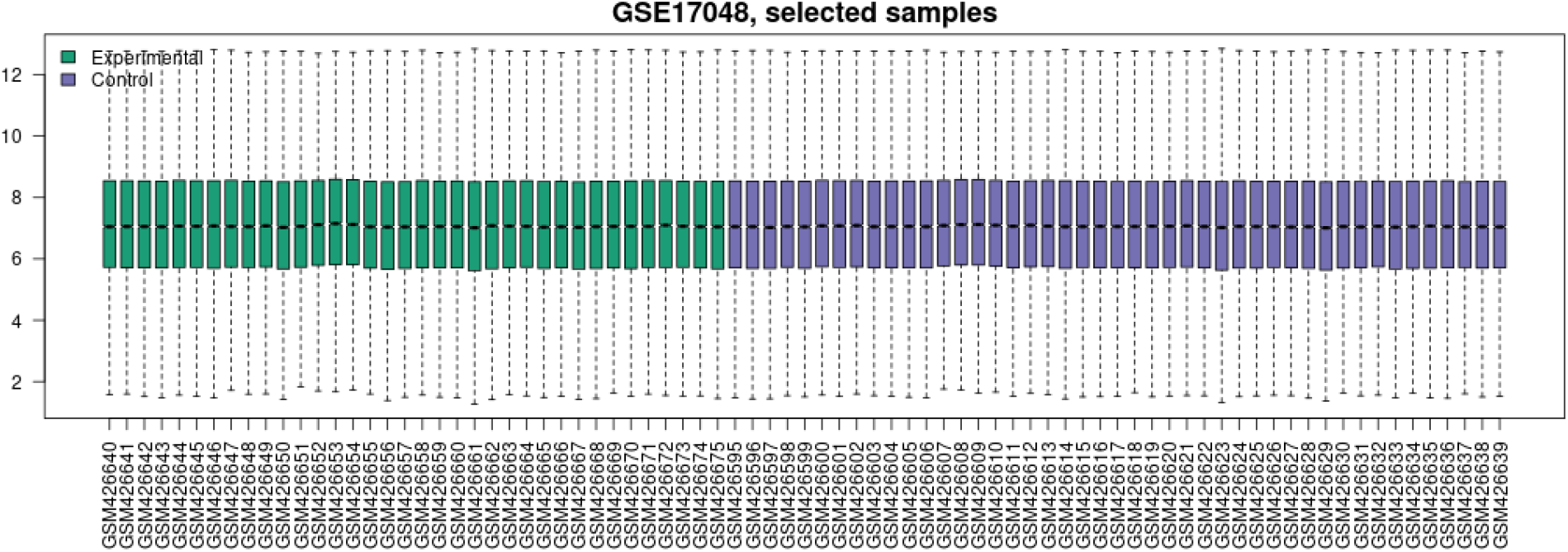
Gene expression value distribution for dataset GSE17048 with each box representing the gene expression values for 1 sample. The green bars correspond to the experimental samples of MS patients while the purple bars correspond to the control samples of the healthy individuals.

**Figure 2 & 3:**
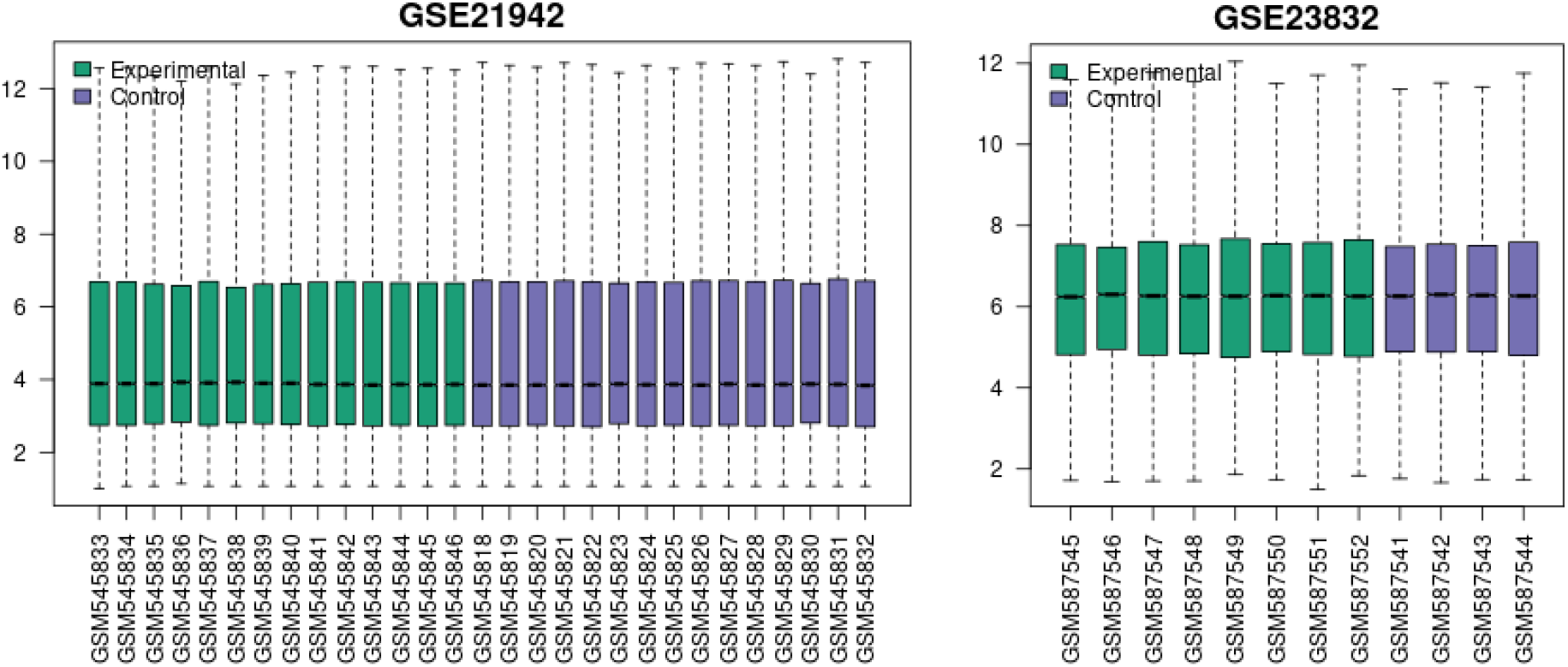
Note “Selected Samples” is not included in the graph title as the entirety of the GSE21942 and GSE2382 datasets was used.

### Identifying Common Genes

The browser-based tool Jvenn was utilized to create a final list of DEGs encompassing the three datasets. If a gene was common to at least 2 out of the 3 datasets, it was kept in the list. Else, it was omitted from the list [14].

### Protein Network Analysis

The final list of DEGs was analyzed in the Search Tool for the Retrieval of Interacting Proteins, STRING, a browser-based tool that scans other databases to create a network map displaying interactions between the protein products of the genes [15]. STRING to construct a network of the protein-protein. STRING also provides statistically significant biological pathways and processes the protein products are involved in. A tab-separated values document containing the data from the network map was downloaded and imported into Cytoscape, an application that can perform network analyses [16]. A table consisting of node degree, clustering coefficient, among other characteristics was generated using Cytoscape’s Network Analyzer Feature. The table generated was then sorted by node degree. Given the distribution of node degrees, all genes with a node degree >= 10 were identified to be hub genes. Hub genes are genes with high levels of interaction with other genes. Since genes with a higher node degree are connected to a greater number of genes, they are more likely to be Hub Genes [17].

### Gene Ontology Analysis

Gene Ontology (GO) is an online database that includes information on biological pathways, processes, components, and the genes that affect them [18-19]. GO’s PANTHER tool takes in as input a list of genes and returns statistically relevant biological processes, pathways, etc. that the genes are involved in [20]. The final list of DEGs was provided as input to PANTHER and molecular pathways and functions were identified in the results.

## Results

**Figure 4:**
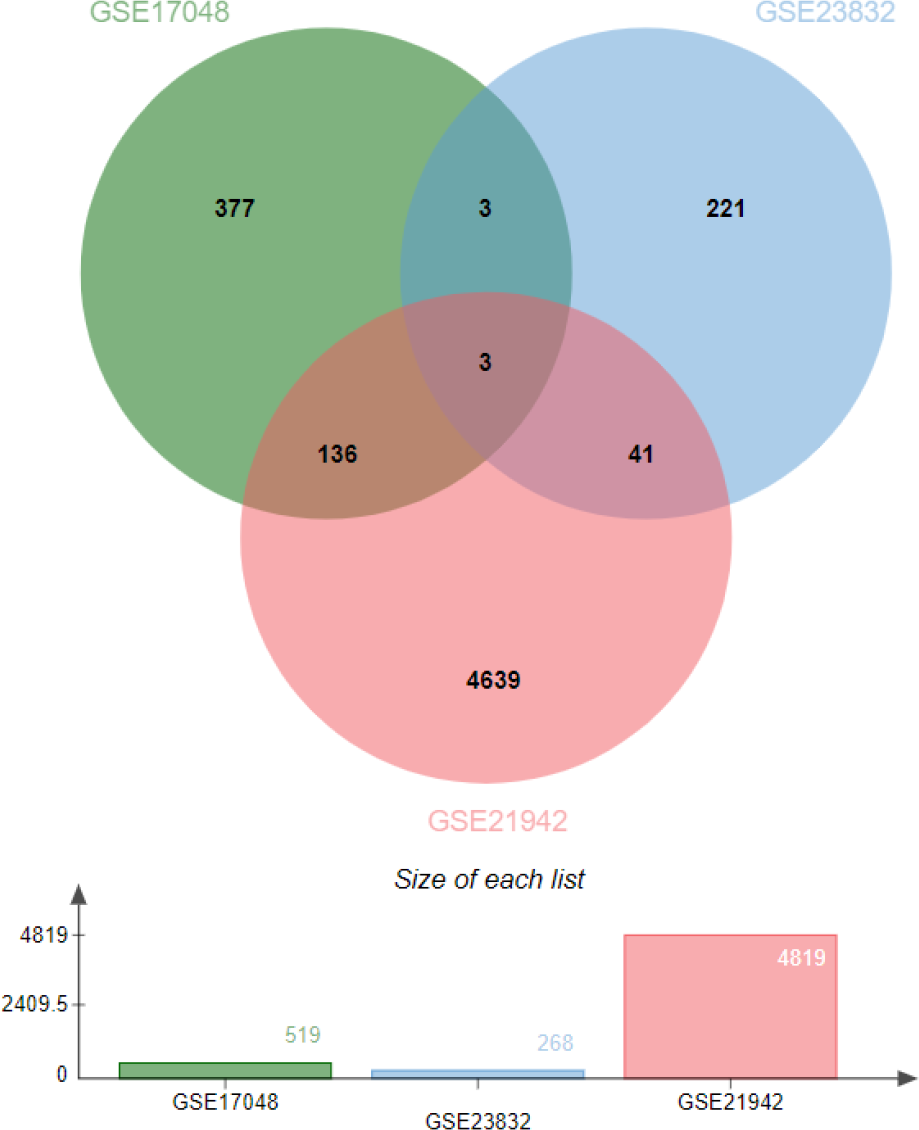
The Venn Diagram depicts the overlap of genes between the 3 datasets. For example, there are 41 genes that are common to GSE23832 and GSE21942.

The three datasets, GSE17048, GSE21942, and GSE23832 were analyzed in GEO2R and the statistically significant DEGs were downloaded and then each set of DEGs (corresponding to the individual datasets) was imported to Google Sheets. During this step, no data was omitted as there were no outliers. Then, the cut-off criteria were set - genes with a p-value (T-Test Results) < 0.05 and fold change value < 0 were removed from the lists. 653 DEGs in GSE17048, 7552 DEGs in GSE21942, and 279 DEGs in GSE23832 passed the criteria requirements. The analysis conducted on Jvenn revealed that 183 of the DEGs were found in at least 2 of the 3 datasets and 3 genes found in all 3 of the data sets. Of these, the two genes coding for significant proteins involved in biological pathways were BRX1 and SCFD1. The BRX1 gene is involved in the biogenesis of ribosomes [21]. The SCFD1 gene is involved in vesicular transport between the ER and Golgi Body [22].

The 183 DEGs that passed the criteria requirements were submitted to the STRING tool. 164 of the selected 183 DEGs were identified and mapped by the STRING tool. The minimum confidence score was set to medium (0.400) by default. To map more accurate protein interactions, the confidence was set to high (0.700).

**Figure 5:**
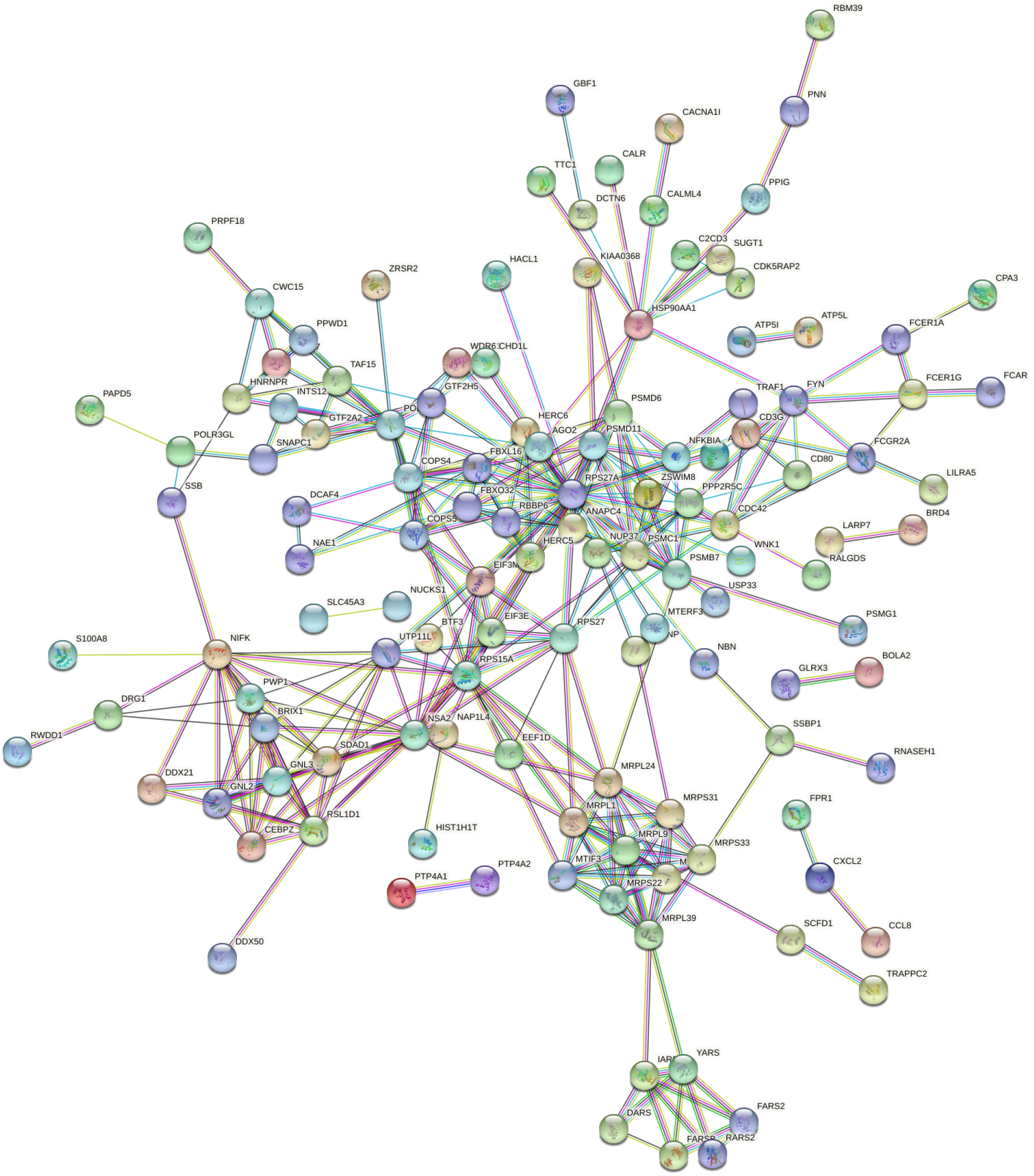
This network generated by STRING displays 164 DEGs as the isolated genes have been omitted. The color of each protein is not of relevance but the lines connecting the nodes indicate how the interaction was determined. For example, the light blue line indicates that information regarding the interaction was already present in the database while the pink line indicates that the interaction was experimentally demonstrated.

STRING also identified some biological processes and molecular functions involving these genes. Some of these include: tRNA aminoacylation for protein translation, immunoglobulin binding, and mitochondrial translation.

The protein interaction network generated on STRING was then downloaded as a TSV and uploaded onto Cytoscape. The genes were sorted on Cytoscape by Node Degree in order to identify the Hub Genes with a minimum Node Degree of 10.

**Figure 5:**
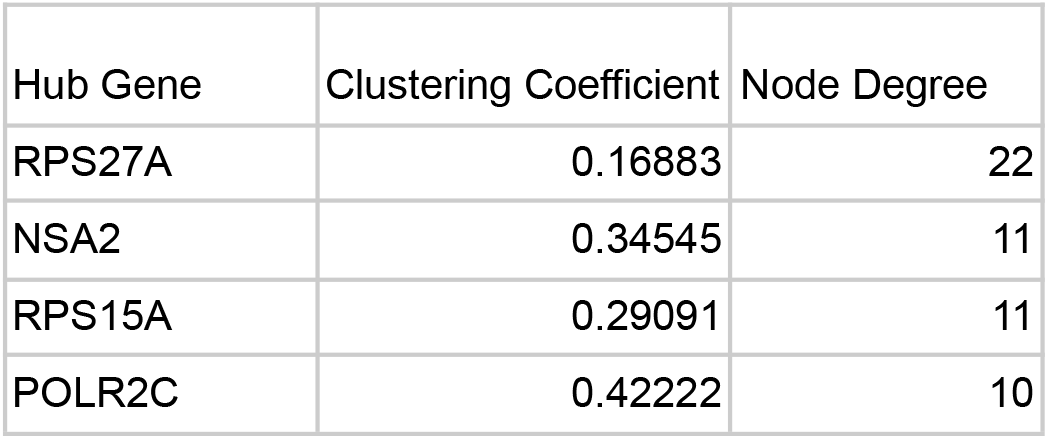
The clustering coefficients and Node Degrees were calculated using Cytoscape’s Network Analyzer Tool. Given the distribution of Node Degrees, all genes with a node degree above 10 were selected as Hub Genes. These include: RPS27A, NSA2, RPS15A, and POLR2C. The local clustering coefficient quantifies how clustered others nodes are around a single node. A clustering coefficient is between 1 and 0 with 1 being the maximum possible density of a cluster around a node and 0 indicating the least density.

RPS27A is a gene that codes for a fusion protein consisting of ubiquitin and a ribosomal protein [23]. Ubiquitins are highly conserved proteins that target other cellular proteins for degradation by Proteasome 26S [24]. In addition to ribosomal functions, it has been seen that RPS27A promoted proliferation, regulated the cell cycle progression, and inhibited the apoptosis of leukemia cells in a past experiment [25]. RPS27A was upregulated in GSE17048 and GSE21942.

Finally, Gene Ontology (GO)’s PANTHER tool was used to analyze the list of DEGs.

The default PANTHER settings were used:

> Test Type: Fisher’s Exact
>
> Correction: Calculate False Discovery Rate

The PANTHER tool was able to identify multiple statistically significant biological processes and molecular functions. The biological process “tRNA aminoacylation for protein translation” was enriched by 10.81 times with a false discovery rate of 1.16E-02 and a p-value of 3.44E-05. The enrichment value is calculated by dividing the number of genes in the provided list of genes that are involved in the biological process by the expected number of genes that would be involved in the process when studying a random sample of genes. Higher enrichment values indicate that larger numbers of genes are involved in a particular Gene Ontology process. Other enriched processes include “mitochondrial translational elongation”, “mitochondrial translational termination”, “Fc receptor signaling pathway”, and “antigen processing and presentation of peptide antigen”.

## Discussion

Multiple Sclerosis is a neurodegenerative disease that is difficult to diagnose at early stages. While there is no cure for the disease, it is advantageous to detect the disease at its onset as early treatment would prevent the rapid progression of the disease and potential disabilities caused by some severe symptoms.

Most of the time, MS can be diagnosed based on visible symptoms and MRI scans. However, sometimes, a spinal tap procedure needs to be performed to substantiate a diagnosis. Because this involves many risks like any other invasive procedure, it is vital to study and identify potential blood-based biomarkers for multiple sclerosis; detection of MS through blood tests would pose much lesser risks, be quicker, and be more affordable.

Analysis of three microarray datasets on MS gene expression yielded 183 differentially overexpressed genes that were common across the studies. These genes passed the p value criteria and warrant further study due to their biological relevancy. While 180 genes were present in two of three studies, only three, BRX1, SCFD1, and C15ORF57 were common to all 3 datasets.

RPS27A, NSA2, RPS15A, and POLR2C all had at least ten interactions with other genes, showing that they are important genes that may be associated with the onset of MS.

RPS27A was identified as a hub gene with a node degree of 22. In addition to ribosomal functions, it has been seen that RPS27A has also been associated with proliferation and regulation of cell cycle progression. It was upregulated in GSE17048 and GSE21942. A study shows that RPS27A may be involved in controlling microglia activation and linked to the onset of neurodegenerative diseases [26]. Like RPS27A, the ribosomal protein gene RPS15A was also identified as a hub gene because it had a node degree of 11.

The NSA2 gene codes for proteins involved in cell cycle regulation and proliferation. In a study investigating TGF beta-inducible nuclear protein 1 (TINP1), a protein encoded for by the NSA2 gene, it was seen that TINP1 overexpression significantly promoted leukemia tumor cell proliferation [27]. NSA2 was upregulated in GSE17048 and GSE21942 and had a node degree of 11.

The POLR2C encodes the third largest subunit of RNA polymerase II. Past studies show that POLR2C mutations have been linked to Primary Ovarian Insufficiency (POI) in Women. It is estimated that anywhere from 4–30% of POI cases are autoimmune in origin [28]. Given the upregulation of the POLR2C in some patients with MS which is an autoimmune disease, the impact of the POLR2C on immune pathways should be studied.

Based on the Gene Ontology Analysis, the most enriched process was “tRNA aminoacylation for protein translation” with an enrichment value of 10.81. Aminoacyl-tRNA synthetases (ARSs) facilitate protein synthesis by linking amino acids to their respective tRNA. Recent studies have shown that these enzymes are also involved with immune cell development (maturation, transcription, activation, and recruitment of immune cells) and immune responses. ARSs have been associated with the development of AASD, an autoimmune disease. Furthermore, studies have shown that ARSs have been dysregulated in patients suffering from Multiple Sclerosis [29]. Given its association with autoimmune diseases, it would be beneficial to study the ARSs molecular pathways linked to the immune system and monitor levels of ARSs and cytokines in MS patients to better understand the relationship and identify whether or not there is a correlation.

The mitochondrial translational processes of elongation and termination were also enriched. In MS patients, dysfunction in mitochondria in lesions has been observed [30]. Furthermore, in the progression of MS, inflammatory mediators, such as cytokines, oxidants, and nitric oxide, are released by microglia. This is believed to result in a malfunction of oxidative metabolism in demyelinated axons.

“The Fc receptor signaling pathway” and “antigen processing and presentation of peptide antigen” were also enriched. These processes are directly linked to immune responses. Furthermore, a past study has observed increased expression of receptors for the Fc part of IgG in MS patients [31]. The Fc receptor is an antibody receptor that allows antigen recognition. MS is associated with an abnormal immune response so it would prove beneficial to study the Fc signaling pathway; since the pathway is involved in immune response to many diseases, though, receptor expression in MS should be compared to that in other diseases to identify if there are any conditions specific to MS.

Some limitations to this study do exist. First off, no dataset normalization was performed. In addition to this, no age range or distinction between female and male samples were factored into the analysis. The progression of MS may differ in people of different age groups and/or men and women and thus, some biomarkers may be effective in detecting MS in some patients but not in others. Currently, the disease is most common in individuals aged between 20 and 40 years. Furthermore, twice as many women are affected as men [1]. Future studies could be done to observe if there are any differences in the progression of the disease in people of different age groups and gender and determine if there are biomarkers that universally tend to be dysregulated in MS patients. Lastly, the blood-brain barrier prevents many molecules in the brain from reaching the bloodstream and thus, doesn’t factor in changes that occur in the cerebrospinal fluid [32].

The list of DEGs identified could aid in the detection of MS through noninvasive tests. They could also serve as targets for treatment procedures. The molecular pathways which were enriched could be studied in greater detail to investigate ways to alleviate symptoms of MS and slow down its progression. The enriched processes of the “Fc receptor signaling pathway”, and “antigen processing and presentation of peptide antigen” should definitely be investigated further because they are directly associated with immune responses. In addition to this, the enrichment of the mitochondrial translational processes should also be studied; establishing a relationship between mitochondrial malfunction and MS could play a significant role in shaping the treatment procedure and may even completely switch its course.

## Data Availability

All data produced in the present work are contained in the manuscript

https://www.ncbi.nlm.nih.gov/geo/query/acc.cgi?acc=GSE17048

https://www.ncbi.nlm.nih.gov/geo/query/acc.cgi?acc=GSE21942

https://www.ncbi.nlm.nih.gov/geo/query/acc.cgi?acc=GSE23832

## Supplemental

List of DEGS:

https://docs.google.com/spreadsheets/d/1a1Y8gFr2x7SuuVqaFqLP-6TGNGzupvdAhluHmtuwsN8/edit?usp=sharing

Venn Diagram Results:

https://docs.google.com/spreadsheets/d/1BQZKqdp2CV3QV5nUEsqSg1ygegLmqRygjlqPdbSxYww9E2M/edit?usp=sharing

StringDB Network:

https://string-db.org/cgi/network?taskId=bYpR9nGy70pi&sessionId=bdkcsENBTGb9

